# A rapid and sensitive method to detect SARS-CoV-2 virus using targeted-mass spectrometry

**DOI:** 10.1101/2020.07.27.20161836

**Authors:** Praveen Singh, Rahul Chakraborty, Robin Marwal, Radhakrishan V S, Akash kumar Bhaskar, Himanshu Vashisht, Mahesh S Dhar, Shalini Pradhan, Gyan Ranjan, Mohamed Imran, Anurag Raj, Uma Sharma, Priyanka Singh, Hemlata Lall, Meena Dutta, Parth Garg, Arjun Ray, Debasis Dash, Sridhar Sivasubbu, Hema Gogia, Preeti Madan, Sandhya Kabra, Sujeet K Singh, Anurag Agrawal, Partha Rakshit, Pramod Kumar, Shantanu Sengupta

## Abstract

In the last few months, there has been a global catastrophic outbreak of severe acute respiratory syndrome disease caused by the novel corona virus SARS-CoV-2 affecting millions of people worldwide. Early diagnosis and isolation is key to contain the rapid spread of the virus. Towards this goal, we report a simple, sensitive and rapid method to detect the virus using a targeted mass spectrometric approach, which can directly detect the presence of virus from naso-oropharyngeal swabs. Using a multiple reaction monitoring we can detect the presence of two peptides specific to SARS-CoV-2 in a 2.3 minute gradient run with 100% specificity and 90.4 % sensitivity when compared to RT-PCR. Importantly, we further show that these peptides could be detected even in the patients who have recovered from the symptoms and have tested negative for the virus by RT-PCR highlighting the sensitivity of the technique. This method has the translational potential of in terms of the rapid diagnostics of symptomatic and asymptomatic COVID-19 and can augment current methods available for diagnosis of SARS-CoV-2.

## Introduction

The world is in the midst of a pandemic caused by severe acute respiratory syndrome corona virus 2 (SARS-CoV-2). More than 15 million individuals from 200 countries have been infected with this virus, which resulted in over 600,000 deaths [1]. Disconcertingly, even after seven months from the first reported case in Wuhan, China, in December, 2019 [2], the number of cases are on the rise with more than 200,000 new cases being reported daily [1].One of the way to reduce the disease propagation is early diagnosis and isolating people infected with the virus. For this, mass testing is the need of the hour. At such a scale, it is critical to innovatively leverage existing infrastructure to build surge capacity and investigate low cost, high throughput methods.

Among the various methods available or developed recently, for diagnosis of SARS-CoV-2, RT-PCR is still the gold standard. Despite RT-PCR being well established and widely available, many problems have emerged in actual performance, ranging from suboptimal sensitivity, especially as the virus mutates, to low throughput due to many intermediate steps and long reaction times[3]. Additionally, specialized reagents are necessary for RT-PCR, which increases the cost of the test. Apart from this, rapid antigen tests are also being extensively used in various countries for preliminary screening. These tests are based on detection of antigens from the nasopharyngeal swab using antibodies. A positive test confirms the presence of the virus, but a negative test is inconclusive since the sensitivity of the antigen testis between 34-80% [4]. Thus, a highly sensitive method is required to directly detect the virus from the nasopharyngeal swab to facilitate rapid screening. Mass spectrometer with high accuracy and sensitivity can be an ideal platform to identify viral peptides with medium to high throughput.

Mass spectrometry (MS) based viral peptide detection have earlier been used for detection of viral proteins [5-7].Identification of SARS-CoV-2 specific peptides from nasopharyngeal swab have also been reported recently [8-10]. Gouveia et al. [8] used SARS-CoV-2 infected cell lysate to identify 2 peptides and validated these peptides in nasopharyngeal swab only in 5 samples with 60% sensitivity. Ihling et al. [9] used gargle solution of 3 infected patients and report the identification of 1 peptide from nucleoprotein using a 180 minute gradient. Cardozo et al. [10] have reported the detection of three peptides using a 10.5 min run time with 83% sensitivity and 96% specificity.

In this study, we report a simple and rapid method that could detect two peptides, QIAPGQTGK and AIVSTIQRKYK, from structural spike glycoprotein and replicase polyprotein 1ab respectively in 2.3 minute gradient with a sensitivity of 90.4% and specificity of 100%. Using follow up samples of patients who have symptomatically recovered and also tested negative for RT-PCR analyses, we show that these peptides are present in these samples indicating the potential of asymptomatic diagnosis of COVID-19 in patients.

## Methods

### Clinical sample collection

Upper respiratory tract sample (nasopharyngeal swab and oropharyngeal swab) were collected in virus transport medium and samples were analyzed using COBAS 6800 automated system (Roche, Basel)according to the manufacturers guidelines[11]. This study was approved by institutional ethics committees of National Centre for Disease Control, New Delhi and CSIR-Institute of Genomics and Integrative Biology, Delhi.

### Sample preparation

Naso and oropharyngeal swab collected in viral transport media (VTM) were inactivated by incubation of samples with lysis buffer (25% guanidinium thiocyanate and 5% SDS) for 20 min at room temperature [12]. To this, 100 µl sodium deoxycholate (0.15% w/v) per ml of sample was added and was incubated for 10 min at room temperature followed by addition of 20% (w/v) trichloroacetic acid (TCA) for protein precipitation. The samples were then centrifuged at 15,000g for 15 min at 4°C. Pellets were washed three times with pre-chilled acetone and protein pellets were resuspended in 100 mM Tris-HCl with 8 M urea (pH-8.5).

### 1. Identification of SARS-CoV-2 proteins using high-resolution mass spectrometry

Proteins extracted from the nasopharyngeal swab of eight RT-PCR confirmed COVID-19 patients were pooled and 300 µg of protein was reduced with 25mM dithiothreitol for 25 minutes at 56°C, followed by alkylation using 55mM iodoacetamide (IAA) at room temperature for 20 minutes in dark. This sample was subjected to trypsin digestion (sequencing grade, Promega) using an enzyme-substrate ratio of 1:10 for 18 hours at 37°C and dried under vacuum. The digested peptides were then fractionated into 8 fractions using cation exchange chromatography using a SCX Cartridge (5 micron, 300 Å bead from AB Sciex, USA).A step-gradient of increasing concentration of ammonium formate buffer (35 mM, 50 mM, 75 mM, 100 mM, 125 mM, 150 mM, 250 mM and 350 mM ammonium formate, 30% v/v acetonitrile and 0.1% formic acid; pH = 2.9) was used to elute the peptides.

### 1. (b). LC-MS/MS data acquisition

The fractionated peptides were analyzed on a quadrupole-TOF hybrid mass spectrometer (TripleTOF 6600, Sciex, USA) coupled to a nano-LC system (Eksigent NanoLC-425) in data dependent acquisition (DDA) mode. Peptides from each fractions were cleaned using C18 Ziptip (Merck, USA) and 4 µg of these peptides were loaded on a trap-column (ChromXP C18CL 5µm 120Å, Eksigent) where desalting was performed with a flow rate of 10 µl per minutes for 10 minutes. Peptides were separated on a reverse phase C18 analytical column (ChromXP C18, 3µm 120 Å, Eksigent) at a flow rate of 5 µl/minute using buffer A (water with 0.1 % formic acid) and buffer B (acetonitrile with 0.1 % formic acid) with following gradient: buffer B was increased gradually from 3% to 25% in first 38 minutes. It was increased to 32% solvent B in next 5 minutes, in the next 2 minutes buffer B was ramped up to 80% and 90% in further 0.5 min. It was held at the same concentration for next 2.5 minutes. Buffer B was brought to initial 3% concentration in next 1 minute and column was reconditioned for 8 minutes before next run.

Ion source parameters were set to 5.5 kV for the ion spray voltage, 25 psi for the curtain gas, 15 psi for the nebulizer gas and 250°C as temperature. For DDA, a 1.8 s instrument cycle was repeated in high sensitivity mode throughout the whole gradient, consisting of a full scan MS spectrum (400–1250 m/z) with accumulation time of 0.25 s, followed by 30 MS/MS experiments (100–1500 m/z) with 0.050 s accumulation time each, on MS precursors with charge state 2+ to 5+ exceeding a 150 cps threshold. Rolling collision energy was used and former target ions were excluded for 15 s.

### 1.(c). Database search

For viral protein identification, a merged search for 8 DDA runs was performed in Protein Pilot software v5.0.1 (Sciex, USA) with paragon algorithm.. The parameters used were as follows: Cysteine alkylation—IAA, digestion—trypsin and 2 missed cleavages were allowed. The search effort was set to ‘Thorough ID’ and false discovery rate (FDR) analysis was enabled. Proteins identified with 1% global FDR were considered. The search was carried out against a UniProt database containing thirteenSARS-CoV-2 proteins.

## 2. Selection of peptides for validation phase

### 2. (a). Protein BLAST search

Protein BLAST search was performed for each of the identified peptide with non-redundant protein sequences (nr) database using NCBI blastp suite by using default parameters for short sequence search.

### 2.(b). Peptide variability analysis

To check whether the identified peptides fall in the invariable region of proteins in SARS-CoV-2, their sequence homology was checked with protein sequences of around 54,000 SARS-CoV-2 strains submitted in the Global Initiative on Sharing All Influenza Data (GISAID) in FASTA format (version 01July2020).Different protein sequences were segregated and aligned, to observe the variability of the identified peptides amongst different strains. JalView tool was used for viewing the consensus of peptides over all the strains [13].

## 3. Detection of SARS-CoV-2 specific proteins

### 3. i. Method standardization

A short scheduled multiple reaction monitoring (sMRM) method, with a retention time window of 20 seconds was developed with the unique viral peptides identified in DDA runs, with multiple transitions for each of the peptides. Sample preparation was performed as described above and data acquisition was performed using a triple quadrupole hybrid ion trap mass spectrometer (QTrap 6500+, Sciex) coupled with a Sciex ExionLC UHPLC system.Tryptic peptides were loaded on an Acquity CSH C18 column (1.7µm 2.1×100mm, Waters) and separated using a binary gradient with buffer A (0.1% formic acid in water) and buffer B (0.1% formic acid in acetonitrile). Peptides were loaded on column with a flow-rate of 600 µL per minute and 98% buffer A. Buffer B concentration was increased from initial 2% to 10% in 0.2minutes and to 50% in 1.1minutes. Buffer B concentration was ramped-up to 90% in another 0.2minute and held at the same gradient for 0.1 minute and then brought to initial 2% in the next 0.1minute and held at the same concentration for 0.6. The total gradient was for 2.3 minutes. Peak review was performed using MultiQuant 3.0(Sciex).

### 3. ii. Serology test for detection of antibody against SARS-CoV-2

To check if the individuals had developed antibodies against SARS-CoV-2, we collected blood samples (5 ml) in vacutainer tubes from individuals and separated the plasma by centrifugation. Presence of antibodies was detected using Elecsys anti-SARS-CoV-2 on Cobas e411, Roche.

## 4. Results

### 4.1. Identification of SARS-CoV-2 proteins using High-resolution Mass Spectrometry

We initially performed a data dependent acquisition of a pool of eight symptomatic RT-PCR confirmed patients using high resolution mass spectrometer as mentioned in the methods section. We identified 22 peptides from 4 proteins of SARS-CoV-2with 1% False discovery rate (FDR). We identified three structural proteins-spike glycoprotein (spike), nucleoprotein (NP) and envelope small membrane protein (ENV) and a protein replicase polyprotein 1ab (Rep) from non-structural part (Supplementary Table 1)-Among these, eight peptides from 3 proteins (Rep, Spike and NP), that were unique and un-modified was selected for generation of an in-house sMRM method for SARS-CoV-2 specific protein detection in patients (Table 1).

**Table 1:** List of unshared and un-modified peptides identified in database search for data dependent run through high-resolution mass spectrometry (HRMS).

| UniProtKB<br>accession<br>no. | Protein name | Peptide sequence | Charge | Parent<br>m/z |
| --- | --- | --- | --- | --- |
| P0DTD1 | Replicase<br>polyprotein<br>1ab | AIVSTIQRKYK | 3 | 436.2662 |
|  |  | LTDNVYIK | 2 | 483.2687 |
|  |  | MDGSIIQFPN | 2 | 561.2684 |
| P0DTC2 | Spike<br>glycoprotein | LIANQFNSAIGK | 2 | 638.3564 |
|  |  | STNLVKNK | 2 | 452.2665 |
|  |  | AHFPREGVFVSNGTHWFVTQR | 4 | 618.8134 |
|  |  | QIAPGQTGK | 2 | 450.2508 |
| P0DTC9 | Nucleoprotein | ADETQALPQR | 2 | 564.7858 |

#### 4.2(a). Protein BLAST search

We also performed Protein BLAST searches with non-redundant protein sequences (nr) database using NCBI blastp suite. The search revealed that seven of the eight peptides were unique to SARS-CoV-2. They were AIVSTIQRKYK and MDGSIIQFPN of Rep, QIAPGQTGK, LIANQFNSAIGK, AHFPREGVFVSNGTHWFVTQR and STNLVKNK of Spike and ADETQALPQR of NP.

#### 4.2(b). Peptides in conserved regions

Sequences for more than 54,000 strains of SARS-CoV-2 have been documented in GISAID up to July 1, 2020. This gave us an opportunity to check if the peptides selected were in the invariant region. Screening through protein sequences of all these strains we found these peptides to be completely conserved among different SARS-CoV2 strains and showed no variability in the peptide sequences (Supplementary Figure 1). Few strain sequences in the GISAID database show gaps and variations, probably due to low sequence quality or host editing.

**Figure 1:**
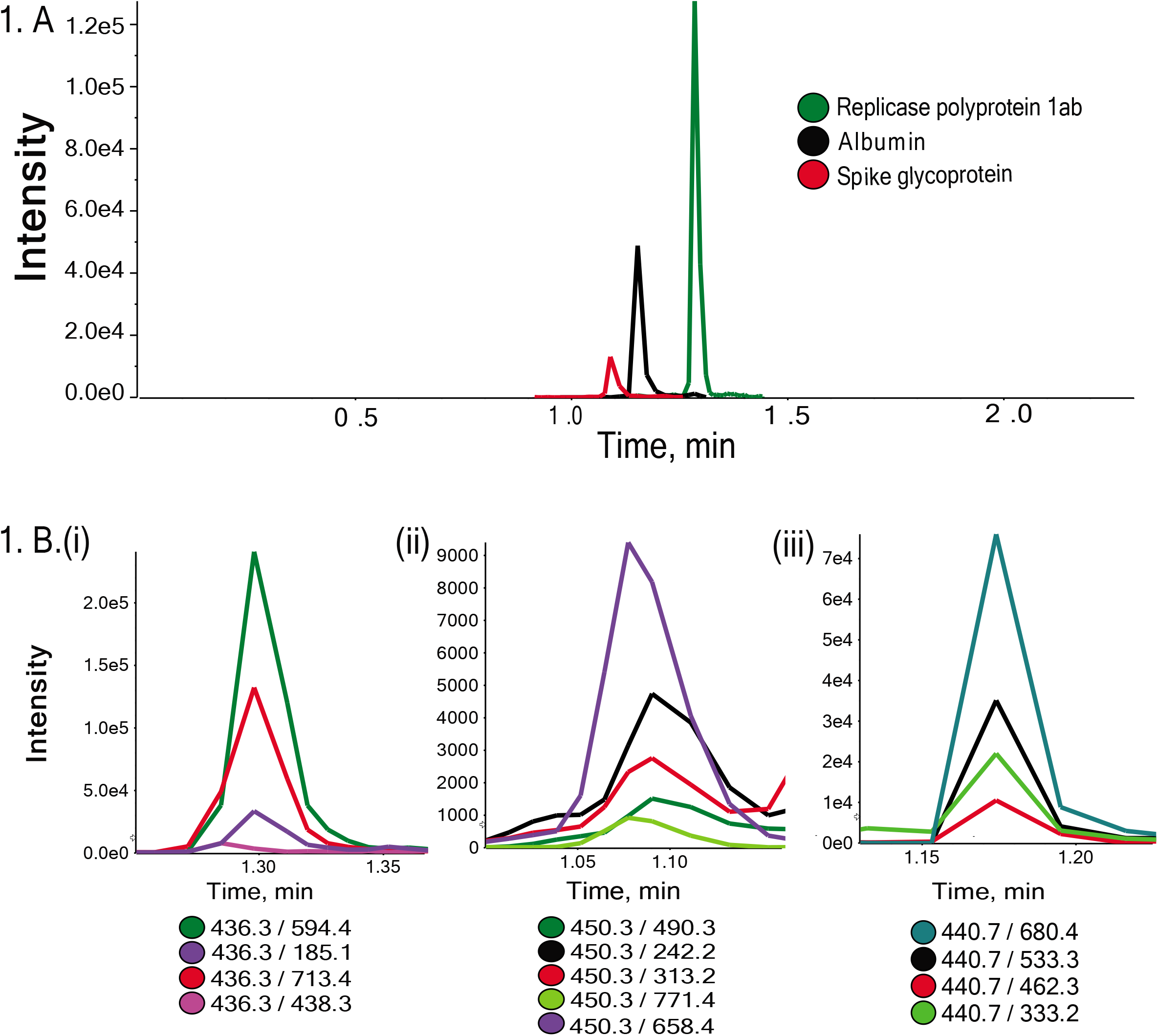
A. Chromatographic separation of the three peptides of naso-oropharyngeal protein digest.Chromatogram for the selected fragment ions are shown in different colours for tryptic digest peptides B.(i) AIVSTIQRKYK (Replicase polyprotein 1 ab), (ii) QIAPGQTGK (Spike glycoprotein) and (iii) AEFAEVSK (Albumin).

#### 4.3. (a) Detection of SARS-CoV-2 specific proteins

For diagnostic purposes, it is important to obtain the desired results in the shortest possible time. Keeping this in mind, we developed a short scheduled multiple reaction monitoring (sMRM) method of 2.3minutes with multiple transitions for each of the identified peptides. Based on the consistency and quality of the peaks in the ion chromatogram we included two peptides-AIVSTIQRKYK from Rep and QIAPGQTGK from spike in the sMRM method (Figure 1). Apart from that, we also included an additional peptide of serum albumin as an internal control for protein amount (Figure 1). The optimized parameters used for the MRM method, to detect SARS-CoV-2 proteins in patient samples are provided in Supplementary Table 2.We considered a sample to be positive for SARS-CoV-2, if overlapping transitions were present with an intensity of greater than 1.5×10^3^ and a signal to noise ratio of greater than 10for either of the two selected peptides.

#### 4.3(b) Detection of SARS-CoV-2 in naso-oropharyngeal swab samples

We analyzed 102 samples using the sMRM method developed in two different sample sets. The first set consisted of 20 samples (including follow up) from 14 patients for whom we had follow up RT-PCR data (Table 2). Interestingly, although most of the patients had recovered as was evident from their being RT-PCR negative and being free of symptoms, we found that they continue to retain the peptides even after 14 days (Sample 5, 6, 7, 8, 10 in Table 2) of initial infection. In 2 of the patients (sample 1 and 14), RT-PCR was found to be negative while the patients had symptoms. One of these patients (sample 14) was found to be negative on the 1^st^ day but was found positive on the second day while we found peptide peaks to be present on both the days, clearly indicating the sensitivity of the sMRM method.

In the second set we analyzed 82 case control samples. Among these 63 were RT-PCR positive and considered as patients. For controls, we considered 19 individuals who had negative RT-PCR and did not have any antibodies as evident from negative serology test indicating the absence of IgM and IgG against SARS-CoV-2. Mass spectrometric analysis revealed that in all the 19 negative samples, peptides were not detected. Of the 63 positive samples, 57 were found to be positive using our method. Thus, our method has a sensitivity of 90.4% and specificity of 100% with respect to RT-PCR positive samples and true controls (serologically negative, RT-PCR negative).

**Table 2:** List of patients considered for set-1 analysis.

| Index patient | Date after 1st RTPCR | Symptoms | RT PCR | Mass Spec |
| --- | --- | --- | --- | --- |
| 1 | 2 | Yes | negative | positive |
| 2 | 2 | No | negative | positive |
| 3 | 3 | yes | positive | positive |
| 4 | 13 | yes | positive | positive |
| 5 | 0 | yes | positive | positive |
|  | 10 | yes | positive | positive |
|  | 18 | No | Negative | positive |
| 6 | 12 | No | Negative | positive |
|  | 16 | No | Negative | positive |
| 7 | 14 | No | Negative | positive |
| 8 | 12 | yes | positive | positive |
|  | 17 | No | Negative | positive |
| 9 | 10 | No | Negative | positive |
| 10 | 10 | No | Negative | positive |
|  | 14 | No | Negative | positive |
| 11 | 10 | No | Negative | positive |
| 12 | 5 | No | Negative | positive |
| 13 | 1 | No | Negative | positive |
| 14 | 0 | yes | Negative | positive |
|  | 1 | yes | positive | positive |

## Discussion

The objective of the study was to develop a simple and rapid targeted mass spectrometric method (sMRM) to identify peptides specific to SARS-CoV-2. For this, we initially analyzed samples in high resolution mass spectrometer and identified eight unique and unmodified peptides from three proteins of SARS-CoV-2. These peptides were unique to SARS-CoV-2 and most importantly lay in the invariant region as revealed by analysis of more than 54,000 SARS-CoV-2 strains sequences submitted in GSAID up to 1^st^ July 2020. This indicates that even if there are mutations in the virus, it should not affect the sequence of the peptides chosen. Using these peptides, we developed a short, 2.3 minutes, gradient for the sMRM method and selected two peptides AIVSTIQRKYK from Replicase and QIAPGQTGK from Spike protein of SARS CoV-2 based on the consistency and quality of the spectra. Apart from these two peptides, we also included one peptide of human serum albumin AEFAEVSK as a control for protein amount. This was important since in a few samples we found that the intensities of albumin transitions were very low indicating low protein concentration and if the viral load is low in such samples it could result in false negative results. These samples were thus not considered.

Using this method we analyzed 102 naso-oropharyngeal samples in two sets. In the first set, we included 20 samples from 14 patients for which RT-PCR data of two or more time points was available. Interestingly, peptide peaks were detected in patients who recovered from the symptoms and were RT-PCR negative for SARS-CoV-2. Interestingly, peptide peaks were observed in two symptomatic patients who tested negative with RT-PCR of which one was tested positive in subsequent RT-PCR indicating false negative RT-PCR in the first instance. Thus, the data indicates high sensitivity of the current method and can be used for initial diagnosis including diagnosis of asymptomatic cases.

In the second set, we analyzed case control samples. As discussed previously, patients who have recovered, and were RT-PCR negative also show peptide peaks. For this, we considered healthy individuals with no exposure to SARS-CoV-2 virus as revealed by serology tests for antibodies against SARS-CoV-2and RT-PCR. These are the true controls since they neither have any active infection, nor had been infected by the virus, previously. We considered a sample to be positive for SARS-CoV-2, if overlapping transitions were present with an intensity of greater than 1.5×10^3^ and a signal to noise ratio of greater than 10 for either of the two selected peptides. This value was chosen since there will be residual peaks from carry-over of the previous sample and it was observed that the median values of the peaks in blank due to carry over were 2×10^2^-4.5×10^2^ depending on the transition with a maximum intensity. Thus, to negate the carry over effect we only considered peaks with intensity more than three times of this (1.5 x10^3^).Using this criterion; we could distinguish SARS-CoV-2 virus infected individuals from uninfected ones with high specificity (100%) and sensitivity (90.4%) when compared with the RT-PCR.

There are several advantages of using mass spectrometer for the diagnosis of SARS-CoV-2. The swab samples can be directly collected in vials containing inactivation buffer ensuring safe transport and handling of the samples. The major advantage of our method is its short run time with a 2.3 minute gradient which is the fastest reported till date for detection of SARS-CoV-2 peptides. Cardozo et al. (2020) reported a MRM with a gradient of 10.5 minutes that could detect SARS-CoV-2 peptides with a sensitivity of 83%. Interestingly, they used fully automated sample preparation protocol using robotic liquid handler and turbulent flow chromatography, multiplexed for online sample clean-up and UPLC separation that enabled them to run 4 samples within the 10.5 min. run. However, this kind of setup will only be available in state-of-the-art laboratories and hence cannot be used for regular diagnostic purpose. In contrast, our method can be used in any laboratory equipped with a triple quadrupole mass spectrometer.

One of the perceived disadvantages of using mass spectrometry based diagnostics is the cost of the equipment, however, the per sample assay cost (less than $3 per sample) is considerably lower when compared to other methods like RT-PCR. We thus, propose that targeted mass spectrometry should currently be explored for screening and diagnostic purpose, which can augment the number of tests that are currently being carried out to detect SARS-CoV-2 and complement traditional RT-PCR based assays or act as an alternate and accurate diagnostic tool with high specificity and sensitivity.

### Data availability

The mass spectometry proteomics data have been deposited to the ProteomeXchange Consortium (http://proteomecentral.proteomexchange.org) via the PRIDE partner repository with the dataset identifier PXD020574.

## Data Availability

The mass spectrometry proteomics data have been deposited to the ProteomeXchange Consortium (http://proteomecentral.proteomexchange.org) via the PRIDE partner repository with the dataset identifier PXD020574.

## Acknowledgement

We thank CSIR project (MLP 2003) for funding. Praveen Singh and Akash kumar Bhaskar received fellowship support from CSIR. Rahul Chakraborty received fellowship from UGC. Anurag Raj received fellowship from DST. Shalini Pradhan, Gyan Ranjan, Mohamed Imran received support from CSIR-IGIB. We thank Purbasha Bhattacharya for helping in writing the manuscript.

## References

1 https://www.worldometers.info/coronavirus

2. Wang C, Horby PW, Hayden FG et al. (2020) A novel coronavirus outbreak of global health concern. Lancet 395:470–473

3. Tahamtan A, Ardebili A (2020) Real-time RT-PCR in COVID-19 detection: issues affecting the results. Expert Rev Mol Diagn 20:453–454

4. Bruning AHL, Leeflang MMG, Vos J et al. (2017) Rapid Tests for Influenza, Respiratory Syncytial Virus, and Other Respiratory Viruses: A Systematic Review and Meta-analysis. Clin. Infect. Dis. 65:1026–1032

5. Majchrzykiewicz-Koehorst JA, Heikens E, Trip H et al. (2015) Rapid and generic identification of influenza A and other respiratory viruses with mass spectrometry. J. Virol. Methods 213:75–83

6. Foster MW, Gerhardt G, Robitaille L et al. (2015) Targeted Proteomics of Human Metapneumovirus in Clinical Samples and Viral Cultures. Anal. Chem. 87:10247–10254

7. Santana WI, Williams TL, Winne EK et al. (2014) Quantification of viral proteins of the avian H7 subtype of influenza virus: an isotope dilution mass spectrometry method applicable for producing more rapid vaccines in the case of an influenza pandemic. Anal. Chem. 86:4088–4095

8. Gouveia D, Miotello G, Gallais F, Gaillard J et al. Proteotyping SARS-CoV-2 virus from nasopharyngeal swabs: a proof-of-concept focused on a 3 min mass spectrometry window. doi:10.1101/2020.06.19.161000. PPR:PPR178294.

9. IIhling C, Tänzler D, Hagemann S et al. Mass Spectrometric Identification of SARS-CoV-2 Proteins from Gargle Solution Samples of COVID-19 Patients. Journal of Proteome Research Article ASAP. DOI: 10.1021/acs.jproteome.0c00280

10. Cardozo KHM, Lebkuchen A, Okai GG et al. Fast and low-cost detection of SARS-CoV-2 peptides by tandem mass spectrometry in clinical samples. doi:10.21203/rs.3.rs-28883/v1. PPR:PPR163832.

11. Corman VM, Landt O, Kaiser M et al. (2020) Detection of 2019 novel coronavirus (2019-nCoV) by real-time RT-PCR. Euro Surveill 25

12. Pastorino B, Touret F, Gilles M, Lamballerie XD, Charrel RN (2020) Evaluation of heating and chemical protocols for inactivating SARS-CoV-2. https://doi.org/10.1101/2020.04.11.036855

13. Waterhouse, A.M., Procter, J.B., Martin, D.M.A, Clamp, M. and Barton, G. J. (2009) “Jalview Version 2 - a multiple sequence alignment editor and analysis workbench” Bioinformatics 25 (9) 1189–1191 DOI: 10.1093/bioinformatics/btp03313.

